# Levels of Person Centered Antenatal Care among Pregnant Women in Public and Private Health Facilities in Western Hararghe Zone, Ethiopia: An Institution Based Comparative Mixed Method Study Design

**DOI:** 10.1101/2024.11.23.24317821

**Authors:** Habtamu Solomon Demeke, Girmatsion Fisseha, Kidanu Gebremariam

## Abstract

**Background:** Person-centered Antenatal Care (PCANC) is a component of quality care that is essential to enable maximize health outcomes. It is believed to be one of the reasons why private health facilities have higher quality than public health facilities. However, in Ethiopia, there is a little study that assesses and compares the level of person-centered antenatal care among pregnant women in public and private health facilities.

**Objectives:** To assess and compare the level of person-centered antenatal care among pregnant women attending antenatal care in public and private health facilities in the Western Hararghe zone, Oromia region, Ethiopia, 2020.

**Methods:** An Institution Based Comparative Mixed Method Study Design was conducted among pregnant women in the Western Hararghe zone, Ethiopia from September 01-October 02, 2020. A multistage stage sampling technique was used to obtain 340 pregnant women (170 from public facilities and 170 from private facilities). The data were entered into Epi-data version 4.4 and analyzed using SPSS version 27. A comparison of categorical and continuous variables was done using an independent t-test and Chi-square test respectively. All comparisons were considered to be significant at a p-value of ≤ 0.05. Qualitative data was analyzed by manually.

**Results:** The overall PCANC percentage mean score was significantly higher in private health facilities (73.7%; 95% CI: 70.5–77.1%) compared to public facilities (59.7%; 95% CI: 56.05–63.5%) (P<0.001). Private facilities had higher percentage mean score than public ones in effective communication (71.4% vs. 56.3%), respect and dignity (85.2% vs. 71.2%), and supportive care (70.1% vs. 59.2%). Qualitative findings further highlighted better interpersonal communication in private facilities.

**Conclusion:** This study highlights significant disparities in the PCANC between private and public health facilities, with private facilities better in effective communication, respect, dignity, and supportive care. Qualitative findings further underscore better interpersonal communication in private settings. Addressing the disparities in person-centered antenatal care between private and public health facilities is crucial to achieve equity and quality of antenatal care.

**Key messages of the article:** *What is already known on this topic:* Person Centered Antenatal Care (PCANC) is critical for ensuring the quality maternal health services. However, disparities in the quality of care contribute to inequities in maternal health outcomes.

*What this study adds:* This is the first study to assess and compare the level of PCANC in private and public health facilities in Ethiopia using the WHO Quality Framework on Maternal and Newborn Care. The study reveals significant inequities in PCANC with private facilities scoring substantially higher in effective communication, respect and dignity, and supportive care compared to public facilities.

*How this study might affect research, practice, or policy:* The findings emphasize the urgent need to address inequities in antenatal care by strengthening Person-Centered care in public health facilities. The findings highlight the need for targeted strategies that enhance communication, respect, and supportive care to ensure equitable and quality antenatal care. Policymakers and health system planners should leverage the WHO framework to implement targeted interventions aimed at improving equity and quality in maternal care services.

## Introduction

Person-centered care (PCC) is care that is respectful and responsive to individual patient preferences, needs, and values, and ensures that patient values guide all clinical decisions ^1^. Even though the term person-centered care has been frequently used in the literature and lacks universal consensus in its definition, this term is used interchangeably with woman-centered care (WCC), respectful maternity care (RMC), patient-centered care (PCC), and client-centered care ^2–7^. The term person-centered care (PCC) is described as the most inclusive term and the most frequently used term in the literature ^3^ ^8–10^. The concepts of person-centered maternity care (PCMC) apply to person-centered antenatal care (PCAC) since antenatal care is one of the key elements of maternity care ^11^ ^12^.

Adapted from the definition by the Person-Centered Care Framework, Person-centered antenatal care (PCANC) refers to providing antenatal care that is respectful of and responsive to individual women and their families’ preferences, needs, and values, and ensuring that their values guide all clinical decisions. It is human rights and is a key domain of quality of care that captures women’s experience of care ^13^. Domains of PCANC include dignity and respect, communication and autonomy, and supportive care, which capture the experience-of-care dimensions in the WHO quality of care framework for maternal and newborn health, and in WHO recommendations for a positive pregnancy experience ^14^ ^15^. These experience dimensions assess person-centered antenatal care (PCANC). A Positive experience during pregnancy is paramount to person-centered care and the human rights of every childbearing woman. In the era of sustainable development goal (SDG), the person-centered antenatal care that brought positive pregnancy experience is so central to achieving the vision to reduce maternal deaths to less than 70 per 100 000 live births by 2030 ^16^.

Person-centered antenatal care is a critical component of quality care to enable treatment adherence and maximize health outcomes ^17^. PCANC has been flagged as a potential strategy for reducing preventable maternal mortality and morbidity to accelerate progress towards meeting the SDG targets for improving maternal and newborn health and a critical component in the continuum of care ^14^ ^18^. PCANC is associated with higher patient satisfaction, earlier presentation for care, improved adherence to post-care treatment, lower healthcare costs having an effective transition to positive labor and birth, and achieving positive motherhood ^19–21^. Poor person-centered care is a driving factor for high maternal mortality, newborn complications, and low rates of facility-based deliveries ^14^ ^22^.

The vast majority of the maternal deaths (94%) occurred in low-and middle-income countries whereby most of these maternal deaths could have been prevented if the women or adolescent girls had been able to obtain person-centered care during pregnancy. Sixty per cent of the stillbirths (1.46 million) occurred during the antepartum period and were mainly attributed to untreated maternal infection, hypertension, and poor fetal growth ^19^ ^23^. If person-centered antenatal care is poor, the women will not attend ANC, irrespective of the number of recommended contacts in the ANC model.

Improving disparities in person-centered antenatal care in public and private health facilities is critical in attaining health equity and the SDG commitment that “no woman is left behind” ^12^ ^24^. Person-centered care is believed to be one of the key reasons why private facilities have higher person-centered maternity scores than public health facilities ^17^. A study conducted in Kenya revealed that the quality of ANC is suboptimal in a dimension of the experience of care whereby those who received care in private facilities had higher experience scores than those who received care in public hospitals PCMC is higher in lower-level and private facilities ^12^. A study conducted in Ethiopia showed that those who received maternity care at private health institutions had significantly higher person-centered maternity care scores when compared with respondents who gave birth in public health institutions ^25^ ^26^.

Despite a paradigm shift at the global level toward person-centered maternity care, assessing person-centered antenatal care has received far less attention ^23^ ^27^. However, some of the studies have focused on person-centered maternity care during childbirth ^25^ ^28^. Very few studies have sought to assess person-centered antenatal care during pregnancy. Moreover, most prior studies on antenatal care have emphasized the service provision dimensions of quality of care by neglecting person-centered antenatal care which captures women’s experience of care during pregnancy ^29–33^. However, little has been said about person-centered antenatal care in public and private health facilities in Ethiopia in general and in West Hararghe. Therefore, the objective of this study was to assess and compare the level of person-centered antenatal care in public and private health facilities in the western Hararghe zone of Ethiopia, 2020.

## Methods

### Study area and period

The study was conducted in the West Hararghe Zone, which lies in the South East part of Ethiopia sharing boundaries with Somali and Afar Regional State to the North, with Bale Zone to the South, East Hararghe Zone to the East, and Arsi Zone and East Showa Zone to the west. West Hararghe Zone is one of the 12 zones in Oromia Regional State. It covers an area of 17,779.4 Km^2^. Available information from the zonal health office shows that the total population of the zone is estimated to be 2,435,350 of which, 2,175,785 (90%) is rural and the remaining 259,565 (10%) is urban. West Hararghe Zone is subdivided into 17 districts whereby there are 11 rural districts of agricultural, 4 rural districts of pastoralist, and 2 urban towns. According to the 2020 zonal health office, the health system of the zone consists of two general hospitals, three primary hospitals, eighty-five health centers, four hundred eighty-two health post facilities, and 25 private clinics. The study was conducted from September 01–October 02, 2020.

### Study Design

An institution-based comparative cross-sectional study triangulated with a qualitative study was used to compare the level of Person-Centered Antenatal care among pregnant women attending antenatal care in public and private health facilities from September 01 –October 02, 2020.

#### Study population

#### Quantitative part

All pregnant women who were attending antenatal care in each of the selected public and private health facilities from September 01–October 02, 2020.

#### For qualitative part

Purposively selected antenatal care providers and antenatal care service users among randomly selected women for exit interviews in public and private health facilities of West Hararghe Zone to assess interpersonal communication attributes of person-centered antenatal care from September 01 –October 02, 2020

##### Inclusion criteria

Pregnant women attending antenatal care in the current facility

##### Exclusion criteria

Women who had lived less than six months in Kebele were severely ill and unable to speak, and those with psychiatric problems, and women who transferred from another health facility to continue their ANC contact were excluded.

### Sample size determination and procedure

The sample size was calculated by using the following double population proportion formula and the Epi Info version ™ 7 software. Using the magnitude of respectful maternity care (P1= 58.1% and P2= 58.1%) from a similar study conducted in Harar, Eastern Ethiopia, with the assumption of 95% Confidence interval, 80% power, design effect of 2 and by adding a 5% non-response rate ^26^. The final sample size estimated was 340, where n1=170 & n2=170.

A multi-stage sampling technique was used; first, seven districts were selected among seventeen districts in the Zone by simple random sampling technique of balloting. The selected districts were Chiro, Hirna, Mieso, Bedessa, Asabot, Mechara, and Gelemso town. Seven health facilities were selected from 14 public health facilities and 7 health facilities from 12 private health facilities were selected using the simple random method from seven districts after identifying public and private health facilities rendering maternity services in seven selected districts. Finally, proportionally allocated sample sizes were made for each selected health facility based on the total number monthly of ANC users in the most recent quarterly report in the selected facility. Individual subjects were selected by systematic random sampling. The K value was calculated by dividing the average monthly ANC attendees by the total sample size of private and public health facilities. That was K=567/170 or 3 for public health facilities and K= 447/170=3 for private health facilities. This interval was used in private and public health facilities separately to select study subjects by systematic random sampling until the required sample size at each health facility based on the order of ANC clients coming to the health facilities. The starting point was a number between 1 and 3 that was selected randomly, which were 2 for both private and public health facilities.

### Qualitative part

#### Interpersonal communication attributes of quality of ANC

One out of three study respondents eligible for client exit interviews was observed by purposive random sampling.

### Measurements

#### Study Variables: Person-Centered Antenatal Care

The PCANC was measured using the PCANC scale which has three domains: effective communication, dignity and respect, autonomy, and supportive care with 6 items of binary response (Yes/No) and 26 items that have a four-point response scale. i.e. 0 (“no, never”), 1 (“yes, a few times”), 2 (“yes, most of the time”) and 3 (“yes, all the time”), and with negative items reverse coded (i.e. questions that were framed negatively, such as the physical, verbal abuse, waiting time, unofficial cost, etc. had to be recorded so that high numbers represent good care. Responses that were recorded as “not applicable” were considered as a missing value. To be consistent with variables with binary responses, we recorded the PCANC variables from a four-point frequency responses scale to binary responses, coding “no, never and a few times” together and “yes, most of the time and all the time” together. As a result, the scale score ranges from **0 to 32** ^34^.

#### Effective Communication scale

Measured using nineteen items with each item has a binary point response scale; i.e. after recoding the PCANC variables from a four-point frequency responses scale to binary responses, 0 (“never or a few times”), 1 (“most of the time or all of the time), so, the total score ranges from 0 to 19 ^34^.

#### Dignity and respect scale

Measured using Six items with a binary point scale; i.e., 0 (“never or a few times”), 1 (“most of the time or all of the time), was used to measure dignity and respect. So, the total score ranges from 0 to 6 ^34^.

#### Supportive care scale

Measured using seven items with each item has a binary point scale; i.e. 0 (“never or a few times”), 1 (“most of the time or all of the time), so, the total score ranges from 0 to 7 ^34^.

#### Socio-demographic variables

Socio-demographic variables of the women include age, residency, educational status, partner’s level of education, religion, marital status, occupation, monthly income, ethnicity, level of participation in household decision-making, domestic violence, self or household member in the facility.

### Qualitative Part

#### Interpersonal communication attributes of Person-Centered Antenatal Care

It includes items related to effective communication with women about the care provided, their; care with respect and preservation of dignity; and access to support care ^18^ ^35^.

##### Data collection tool and procedures

Quantitative Data were collected through client exit interviews by using a pre-tested structured Local Afan Oromo version questionnaire. The study used a standard validated tool adapted from a person-centered maternity care scale developed in Kenya with Cronbach’s alpha of 0.86 ^34^. The other included questions in the questionnaire were prepared by reviewing different works of literature ^12^. The questionnaire in the English and local Afan Oromo versions is available in **Supplementary File 1** and **Supplementary File 2** respectively.

Qualitative data were collected using a semi-structured observation checklist to assess interpersonal attributes of antenatal care as per the WHO quality framework and after reviewing the literature ^18^ ^35^. The questionnaire was first developed in English and translated into the local language Afan Oromo and re-translated back into English by blind translators to ensure its consistency. Data collectors were ten health science University students residing where the study takes place. The training was given to collectors for two days before the normal data collection time, a pretest was conducted using a 5% (17) sample size on a non-selected health facility (Micheta Health Center) and the necessary correction was made accordingly.

All respondents were informed about the purpose of the study, and their written consent was obtained. The respondents’ right to refuse or withdraw from participating in the interview was fully maintained and the information provided by each respondent was kept strictly confidential. For the qualitative part, the investigator who did not participate in the caregiving process observed the provider interaction to assess interpersonal communication attributes of the person-centered antenatal care. Qualitative data was collected and analyzed by the primary investigator. For the observation part of the qualitative study, client-provider interactions on the first three observations were not considered for analysis before actual data collection to minimize the Hawthorne effect. Finally, qualitative findings were categorized to triangulate the quantitative findings.

Internal reliability for this study was checked by calculating Cronbach’s alpha for each of the domains. The PCANC scale used in this study had a maximum possible score of 32. These items were grouped into three domains per WHO’s latest quality framework, comprising effective communication, respect and dignity, and supportive care domains. These subscales have maximum possible scores of 19, 6, and 7 respectively. The Cronbach alpha for the overall PCANC scale was 0.931. For the sub-scale, the reliability test conducted shows that the effective communication scale, respect and dignity scale, and supportive care scale had Cronbach alpha values of 0.889, 0.662 and 0.754 respectively.

##### Data processing and analysis

Data were checked for completeness and consistency, and then coded and entered into Epi. Info version ™ 7 software and exported to SPSS version 27 software package for analysis. A comparison of independent categorical variables was done using the Chi-square test while an independent t-test was used for continuous variables. All comparisons were considered to be significant at a p-value of ≤ 0.05. The descriptive data were analyzed using frequency distribution, percentages, and cross-tabulation. For qualitative data, descriptive qualitative analysis was made manually. Finally, the findings were compiled and presented using tables, graphs, figures, and texts.

##### Patient and public involvement

Patients and /or the public were not involved in the design, conduct, reporting and dissemination of this study.

## Results

### Socio-Demographic Characteristics of Respondents

In this study, a total of 340 women were involved for the study, one hundred seventy from each facility with a response rate of 100%. The details are shown in **Table 1** below.

**Table 1.** Socio-demographic characteristics of respondents in private and public health facilities in West Hararghe zone, Oromia, Eastern Ethiopia, 2020 (n = 340)

| Variables | Category | Private | Public | X<sup>2</sup> (P -value) |
| --- | --- | --- | --- | --- |
| **Age** | Below 19 | 11(6.5%) | 10 (5.9%) | 1.93 (0.75) |
|  | 20-24 | 37 (21.8%) | 42 (24.7%) |  |
|  | 25-29 | 78 (45.9%) | 84 (49.4%) |  |
|  | 30-34 | 38 (22.4%) | 30 (17.6%) |  |
|  | above 35 | 6 (3.5%) | 4 (2.4%) |  |
| **Religion** | Orthodox | 52 (30.6%) | 57 (33.5%) | 1.1 (0.9) |
|  | Muslim | 86 (50.6%) | 84 (49.4%) |  |
|  | Protestant | 23 (13.5%) | 18 (10.6%) |  |
|  | Catholic | 8 (4.7%) | 10 (5.9%) |  |
|  | Other | 1 (0.6%) | 1 (0.6%) |  |
| **Place of residence** | Urban | 111 (65.3%) | 66 (38.8%) | 22.8 (**0.00**<sup>*</sup>) |
|  | Rural | 59 (34.7%) | 104 (61.2%) |  |
| **Level of education** | Illiterate | 12 (7.1%) | 25 (14.7%) | 5.1 (**0.02**<sup>*</sup>) |
|  | Literate | 158 (92.9%) | 145 (85.3%) |  |
| **Partner level of education** | Illiterate | 9 (5.3%) | 33 (19.4%) | 15.6 (**0.00**<sup>*</sup>) |
|  | Literate | 161 (94.7%) | 137 (80.6%) |  |
| **Occupation** | House wife | 62 (36.5%) | 75 (44.1%) | 32.7(**0.00**<sup>*</sup>) |
|  | Merchant | 41 (24.1%) | 35 (20.6%) |  |
|  | Government | 41 (24.1%) | 8 (4.7%) |  |
|  | Farmer | 16 (9.4%) | 34 (20.0%) |  |
|  | Other | 10 (5.9%) | 18 (10.6%) |  |
| **Marital status** | Married | 169 (99.4%) | 170 (100.0%) | 1.000 |
|  | Other | 1 (0.6%) | 0 (0.0%) |  |
| **Ethnicity** | Oromo | 124 (72.9%) | 139 (81.8%) | 5.94 (0.11) |
|  | Amhara | 30 (17.1%) | 16 (9.4%) |  |
|  | Somale | 4 (2.9%) | 6 (3.5%) |  |
|  | Gurage | 12 (7.1%) | 9 (5.3%) |  |
| **Monthly income** | ≤3000 | 69 (40.6%) | 106 (62.4%) | 15.3 (**0.00**<sup>*</sup>) |
|  | >3000 | 101 (59.4%) | 64 (37.6%) |  |
| **Self or household member** | Yes | 11 (6.5%) | 13 (7.6%) | 0.04 (0.83) |
|  | No | 159 (93.5%) | 157 (92.4%) |  |
| **Domestic Violence** | Yes | 49 (28.8%) | 76 (44.7%) | 8.55(**0.03**<sup>*</sup>) |
|  | No | 121 (71.2%) | 94 (55.3%) |  |
| **Participation in household** | Low | 59 (34.7%) | 92 (54.1%) | 12.2(**0.00**<sup>*</sup>) |
|  | High | 111 (65.3%) | 78 (45.9%) |  |

### Level of Person-Centered Antenatal Care

The mean PCANC scale of the respondents in both private and public health facilities was 21.34. In particular, the mean of overall PCANC scale of respondents in private health facilities was 23.58 compared to 19.11 for the respondents at public health facilities (P=0.001, t-test=5.38). Regarding the mean of PCANC subscales, the mean of effective communication subscale of the respondents in both private and public health facilities was 12.13. The mean of effective communication subscale of the respondents in private health facilities was 13.56 while those who had ANC contact at public health facilities had a mean score of 10.7 (P=0.000, t-test=5.52). The mean of respect and dignity subscale of the respondents in both private and public health facilities was 4.69. Specifically, the mean of respect and dignity subscale of the respondents in private health facilities was 5.11 while it was 4.27 for respondents at public health facilities (P=0.000, t-test=5.85). Furthermore, the mean of supportive scale of the respondents in both private and public health facilities was 4.53. The mean of supportive care subscale of the private clients was 4.91 while that of public clients was 4.15 (P=0.001, t-test=3.48).

#### Percentage Mean Score of overall Person-Centered Antenatal Care and its Domains score

The following formula was used to standardize the mean score;

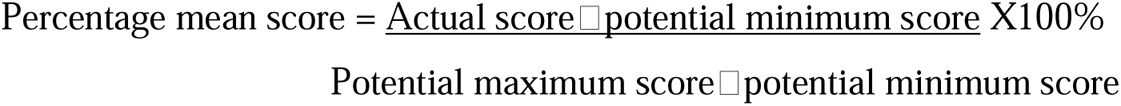

Based on this formula, the percentage mean score of overall PCANC and its domain score is presented in **Supplementary Figure 1.**

### Distribution of Individual Person Centered Antenatal Care items among women in private and public health facilities

#### Effective communication

Most of the respondents 124 (72.9%) who had antenatal contact at public health facilities reported that providers introduce themselves a few times or never while nearly half 84 (49.4%) of the respondents experienced the same at private health facilities (P=0.00). The majority of the respondents 156 (91.8%) at private health facilities were told about the signs of pregnancy complications compared to 123 (72.4%) of the respondents at public health facilities (P=0.000). About 96 (56.5%) of the women respondents at private health facilities understood the purposes of tests performed most or all of the time whereas at public health facilities, 103 (60.6%) of women understood the purposes of tests performed a few times or never (P=0.002). The details of the distribution of effective communications domain are shown in Table 2

**Table 2.**
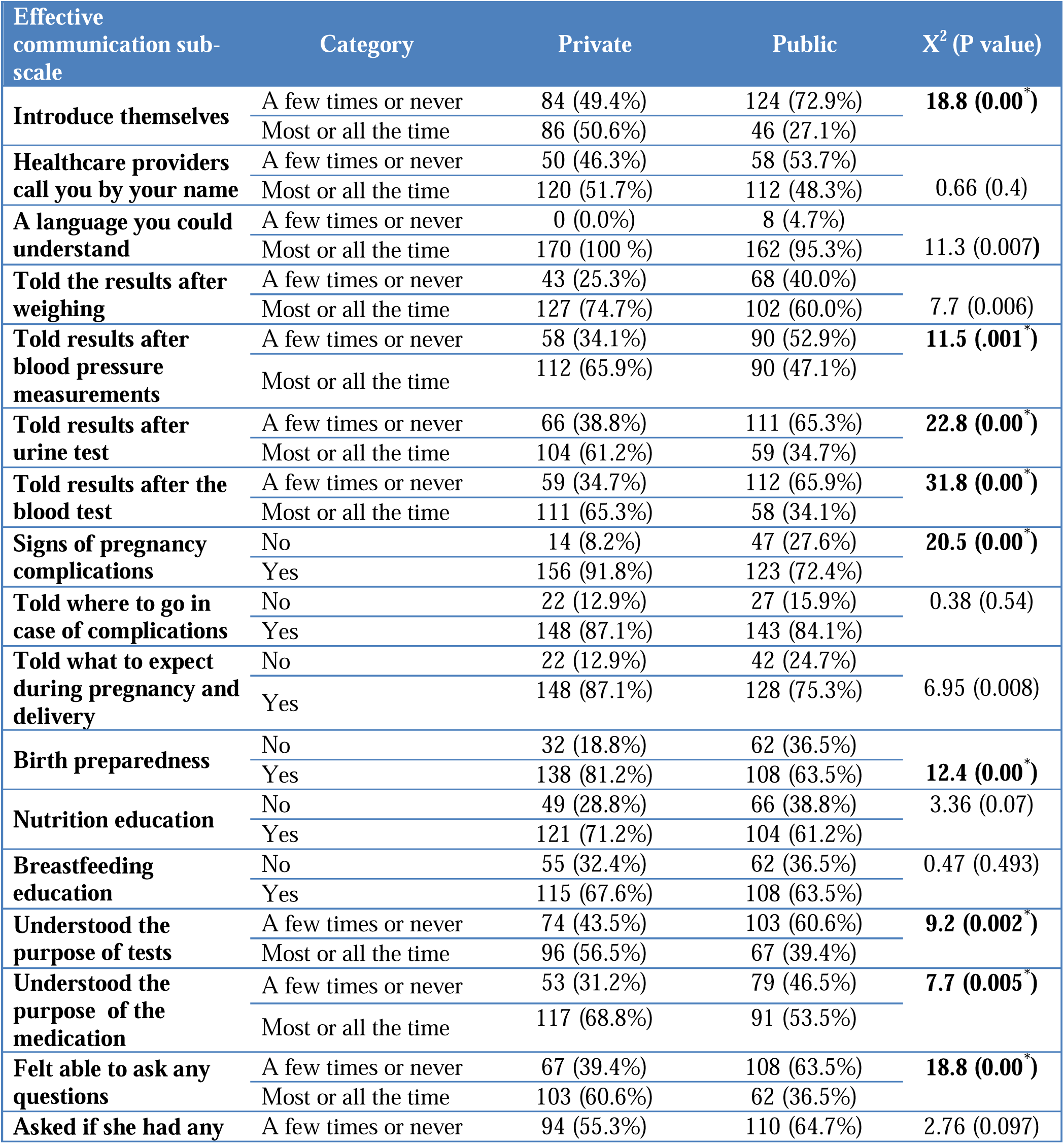
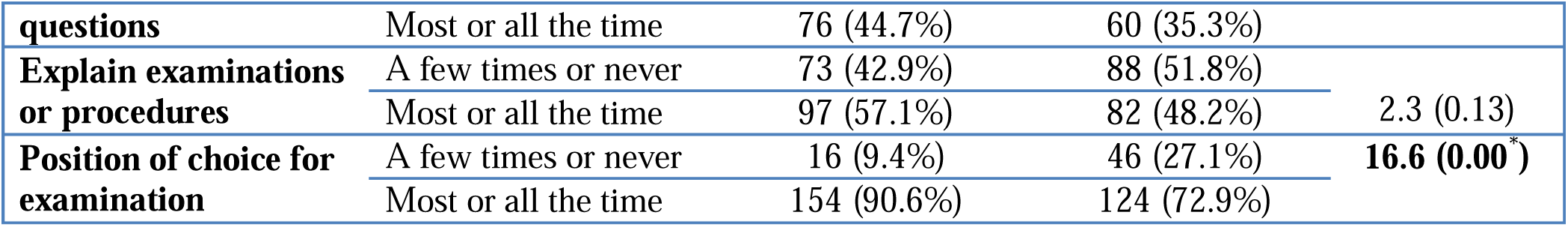
Distribution of effective communications domain among women in private and public health facilities (n = 340), West Hararghe Zone, Ethiopia, 2020.

Table 2 Distribution of effective communications domain among women in private and public health facilities (n = 340), West Hararghe Zone, Ethiopia, 2020

#### Respect and dignity

About more than nine in ten (91.8%) of the respondents at private health facilities felt they were treated with respect most or all of the time while over two-thirds (67.6%) did experience the same for the women at public health facilities (P=0.00). One hundred forty (82.4%) of private clients reported they did not feel exposed most or all the time compared to more than two-thirds of public health facility clients 114 (67.1%) who didn’t feel similar most or all the time (P=0.002). About 7 in 10 of the study subjects 120 (70.6%) at private health facilities felt that they could discuss issues in a private most or all of the time on the other hand more than half of the respondents 88 (51.8%) at private health facilities felt that they could discuss issues in a private a few times or never (P=0.000). The details are shown in **Table 3** below.

**Table 3.** Distribution of respect and dignity domain among women in private and public health facilities (n = 340), West Hararghe Zone, Oromia Region, Eastern Ethiopia, 2020.

| Respect and dignity sub-scale | Category | Private | Public | X<sup>2</sup> (P value) |
| --- | --- | --- | --- | --- |
| **Treat you with respect** | A few times or never | 14 (8.2%) | 55 (32.4%) | **29.1 (0.00\*)** |
|  | Most or all the time | 156 (91.8%) | 115 (67.6%) |  |
| **Treated friendly** | A few times or never | 50 (29.4%) | 93 (54.7%) | **21.2 (0.00\*)** |
|  | Most or all the time | 120 (70.6%) | 77 (45.3%) |  |
| **Health providers shouted scolded, insulted, and threatened at you** | Most or all the time | 2 (1.2%) | 0 (0.0%) | 2.8 (0.5) |
|  | A few times or never | 168 (98.8%) | 170 (100 %) |  |
| **Treated roughly like pushed, beaten etc.** | Most or all the time | 5 (2.9%) | 3 (1.8%) | 0.52 (0.72) |
|  | A few times or never | 165 (97.1%) | 167 (98.2%) |  |
| **Privacy** | A few times or never | 30 (17.6%) | 56(32.9%) | **9.73 (0.002\*)** |
|  | Most or all the time | 140 (82.4%) | 114 (67.1%) |  |
| **Confidentiality** | A few times or never | 50 (29.4%) | 88 (51.8%) | **16.7 (0.00\*)** |
|  | Most or all the time | 120 (70.6%) | 82(48.2%) |  |

#### Supportive care

About 110 (64.7%) of the respondents at private health facilities reported that providers talked to them about their feelings most or all of the time, whereas just half of the respondents 85 (50.0%) at public health facilities reported that providers talked to them about their feelings a few times or never (P=0.008). Nearly half of the private clients 83 (48.8%) reported that they were paid unofficial (additional cost) a few times or many times compared to only 14 (8.2%) of public clients reported that they paid similar a few times or many times (P= 0.00). About 127 (74.7%) of the respondents at private health facilities felt that the waiting time was somewhat short or somewhat short, in contrast to 100 (58.8%) of women who felt the waiting time was somewhat long or very long for the respondents at public health facilities (P= 0.00). The details are shown in Table 4 below.

**Table 4.** Distribution of supportive care domain among women in private and public health facilities (n = 340), West Hararghe Zone, Oromia Region, Eastern Ethiopia, 2020.

| Supportive sub scale | Category | Private | Public | X<sup>2</sup>(P value) |
| --- | --- | --- | --- | --- |
| **Feeling** | A few times or never | 60 (35.3%) | 110 (50.0%) | 6.93 (0.008) |
|  | Most or all the time | 110 (64.7%) | 85 (50.0%) |  |
| **Encourage male involvement** | A few times or never | 72 (42.4%) | 91 (53.5%) | **3.82 (0.05\*)** |
|  | Most or all the time | 98 (57.6%) | 79(46.5%) |  |
| **Unofficial cost** | A few times or many times | 83 (48.8%) | 14 (8.2%) | **66.7 (0.00\*)** |
|  | Never or yes, once | 87 (51.2%) | 156 (91.8%) |  |
| **Treated differently because of any personal attribute** | A few times or many times | 5 (2.9%) | 8 (4.7%) | 0.32 (0.57) |
|  | Never or yes, once | 165 (97.1%) | 162 (95.3%) |  |
| **Felt the health facility was clean** | A few times or never | 46 (27.1%) | 93 (54.7%) | **25.75 (0.00\*)** |
|  | Most or all the time | 124 (72.9%) | 77 (45.3%) |  |
| **Waiting time** | Somewhat long or very long | 43 (25.3%) | 100 (58.8%) | **37.85 (0.00\*)** |
|  | Short or somewhat short | 127 (74.7%) | 70 (41.2%) |  |
| **Felt safe** | A few times or never | 48 (28.2%) | 93 (54.7%) | **23.4 (0.00\*)** |
|  | Most or all the time | 122 (71.8%) | 77(45.3%) |  |

### Qualitative findings

#### Interpersonal Communications Attributes of Person-Centered Antenatal Care

The finding of direct non-participant observation for ten women attending ANC at private health facilities suggested that most of the women were comfortable being provided a seat and greeted by health providers. Apart from this, there was no interruption of speech by the provider and clients were treated respectfully in most of the observations taking place at private facilities. During observation private health facilities, explaining procedures or medical information to the women, allocating sufficient time to clients and examining women in the private room were offered for most of the women from ten observations by observer. Meanwhile, the concerns of women were also asked for some of the clients during observation of private health facilities.

On the other hand, the observation of twenty clients at public health facilities demonstrated that welcoming the patient, providing seats and greeting women to be comfortable were offered for only a few women. Additionally, there was interruption of speech by health providers, clients weren’t treated respectfully for the majority of clients during the time of observation in this study. Explaining procedures or medical information to the women, allocating sufficient time to clients and examining women in private rooms or privacy was offered for a few of the women. Moreover, concerns of women were asked about for few clients attending public health facilities.

## Discussion

This study is the first study that assesses and compares the level of Person-Centered Antenatal Care among pregnant women in public and private health facilities in Western Hararghe zone, Oromia, Ethiopia, 2020. The Percentage Mean score of the overall PCANC scale in both private and public health facilities was 66.7% [95% CI: 64.2, 69.4%]. The domains of effective communication had relatively low Percentage Mean scores followed by supportive care then respect, and dignity. This study revealed significant differences in the level of PCANC between private and public health facilities, with private facilities performing better across domains of effective communication, respect and dignity, and supportive care (p<0.001). Meanwhile, the qualitative study findings reported the interpersonal communication attributes of PCANC at private health facilities were better than the public ones.

The Percentage Mean score of overall PCANC in both private and public health facilities was 66.7% [95% CI: 64.2, 69.4%]. This is in studies consistent with the percentage mean score of the Person-centered Maternity Care (PCMC) scale conducted in Northeastern, Ethiopia (64.5%), urban Kenya (66.9 %) but higher than study conducted in rural Ghana 51.6 % ^25^ ^28^. This implies the form of antenatal care that was rendered to the pregnant mother in these settings was not taking into account women’s preferences, needs, and aspirations. The consequences of this kind of care which is not person-centered care not only deprives or violates human rights it has also a devastating impact on maternal and newborn outcomes ^19^ ^36^ ^37^.

In the domains of PCNC, the Percentage Mean score at both private and public health facilities was 78.2% [95% CI: 75.8-80.5%] for respect and dignity, 63.8 [95% CI: 61.1, 66.8%] for effective communication, and 64.7% [95% CI: 61.8-67.7%] for supportive care. This indicates domains of effective communication had relatively low scores followed by supportive care then respect, and dignity. This finding is consistent with the studies than the study conducted in Northeastern Ethiopia, Enugu Nigeria, and low and middle-income countries ^25^ ^38^ ^39^.

The percentage means score of the overall PCANC scale at private health facilities was 73.7% [95% CI: 70.5, 77.1%] while it was 59.7% [95% CI: 56.05, 63.5%] for respondents at public health facilities (P<0.001). This variation was supplemented by qualitative findings which suggested the interpersonal communication attributes of PCANC at private health facilities were better than the public one. This is consistent with a study reported that women who delivered in private facilities reported a higher PCMC score than those who delivered in public hospitals ^28^. This implied that there are inequities of PCANC in private and public health facilities. This eventually results in divergence within groups, and societies that hampered universal health coverage to achieve SDG goals with the slogan of “NO ONE IS LEFT BEHIND” ^14^ ^16^ ^40^.

In the domain of effective communication, the percentage mean score of PCANC at private health facilities was 71.4% [95% CI: 68.1, 75.01%] compared to public health facilities 56.3% [95% CI: 52.5, 60.4%] (P<0.001). This is supplemented by This was also supplemented by the qualitative study by observation suggesting that most of the women were more effectively communicated by health providers in private facilities than those at public facilities. This implies there is a poor rapport between healthcare providers and women, who attended ANC which affects women’s adherence to treatments and recommendations, willingness to seek care in the future in the facility, and user satisfaction and improve maternal and newborn outcomes ^19^ ^21^.

In the domain of respect and dignity, the percentage mean score of PCANC at private health facilities was 85.2% [95% CI: 82.5, 87.8%] compared to public health facilities 71.2% [95% CI: 67.3, 74.9%] (P<0.001). This finding is supplemented by the qualitative study by the observation that clients were treated respectfully in most of the observations that took place at private facilities while in public health facilities, only a few women were respected. This implies that in about 6 in 10 women, violation of article 4 which protects Women’s right to be treated with dignity and respect is violated ^41^.

In the domain of supportive care, the percentage mean score of PCANC at private health facilities was 70.1% [95% CI: 65.7, 74.6%] compared to public health facilities 59.2% [95% CI: 55.05-63.6%] (P<0.001). This finding is also supplemented by a qualitative study whereby explaining procedures or medical information to the women, allocating sufficient time to clients, and examining women in private rooms or privacy was offered and concerns of women were asked for a few of the women attending public health facilities compared to private one. This indicates the lack of continuous support during pregnancy that can result in low rates of facility-based deliveries thereby increasing the vulnerability of mothers to develop maternal and newborn complications ^14^ ^16^.

### Strengths and limitations

This is the first study that contributes to the growing body of evidence on PCANC providing critical insights into facility-level disparities. The study also employed a comparative mixed-method study design that combined qualitative study to triangulate quantitative findings. The data were collected using systematic random sampling and thereby findings can be more generalized to the target population. Comparing the counts of PCANC domains using the WHO quality framework captures the diversity of person-centered antenatal care that would not have been possible by simple prevalence measures.

Despite these strengths, this study could suffer from social desirability bias as women may not want to report negative experiences when interviewed at the facility due to fear of retaliation. Another potential limitation is the risk of a potential Hawthorne effect despite strong efforts to minimize the bias. This study didn’t identify factors affecting Person Centered Antenatal Care. Future researchers should conduct a study on person-centered Antenatal Care and provisions of care dimensions and underlying factors with structural attributes of care as cross-cutting issues.

## Conclusion

The findings of this study highlight significant disparities in the PCANC between private and public health facilities. Across the three domains, effective communication, respect and dignity, and supportive care, private health facilities consistently achieved higher percentage mean scores compared to their public counterparts. Meanwhile, the qualitative finding suggested that interpersonal communication attributes of PCANC at private health facilities were better than the public ones. Addressing the disparities in person-centered antenatal care between private and public health facilities is critical to attaining health equity and SDG commitment, particularly in resource-limited settings. Prioritizing the allocation of resources to reduce patient-provider ratios, enhance training programs focusing on communication and interpersonal skills, and implement accountability mechanisms to ensure adherence to standards of person-centered care. Additionally, promoting public-private partnerships could facilitate the exchange of best practices and innovations to achieve equity and quality of antenatal care.

## Data Availability

The datasets used for this study are available from the corresponding author upon reasonable request. All supplementary information files are included in this manuscript.

## Acknowledgements

We are grateful to the study participants. We are also grateful for Mekelle University and Samara University, Ethiopia.

## Authors’ contributions

HS, GF and KG had made substantial contributions to the conception and design of the survey. HS has designed the study, participated in data collection, analysis, interpretation, and write-up, drafted the manuscript and critically revised it. GF has participated in study design, analysis, interpretation, and critically revised the manuscript. MK has participated in the study design, analysis, and interpretation drafted the manuscript and critically revised the manuscript. All authors read and approved the final manuscript.

## Ethical approval and consent to participate

The study protocol was reviewed and approved by the ethical review committee of Mekelle University, College of Health Sciences. The expedited approval of this study was received from the ethical review of committee Mekelle University, College of Health Sciences with code **1504/2020**. All the study participants were informed about the purpose of the study and finally, verbal consent was obtained before the interview. Respondents were informed about the purpose of the study, and their oral consent was obtained. The respondent’s right to refuse or withdraw from participating in the interview was fully maintained. Confidentiality and anonymity were ensured.

## Consent for publication

Informed consent was obtained from each subject so that the study would be published and presented at different workshops without the anonymity of the participants.

## Funding and Disclaimer

The study was financially supported by Samara University, the Department of Public Health, and the College of Medical and Health Science. Its contents are solely the responsibility of the authors and do not necessarily represent the official views of the supporting offices. The funders had no role in the study design, data collection, and analysis, decision to publish, or preparation of the manuscript.

## Provenance and peer review

Not commissioned; externally peer-reviewed.

## Supplemental material

This content has been supplied by the author(s). It has not been vetted by BMJ Publishing Group Limited (BMJ) and may not have been peer-reviewed. Any opinions or recommendations discussed are solely those of the author(s) and are not endorsed by BMJ. BMJ disclaims all liability and responsibility arising from any reliance placed on the content.

## Open access

This is an open-access article distributed in accordance with the Creative Commons Attribution Non-Commercial (CC BY-NC 4.0) license, which permits others to distribute, remix, adapt, build upon this work non-commercially, and license their derivative works on different terms, provided the original work is properly cited, appropriate credit is given, any changes made indicated, and the use is non-commercial. See: http://creativecommons.org/licenses/by-nc/4.0/.

## Supplementary Figure 1

**Figure 1.**
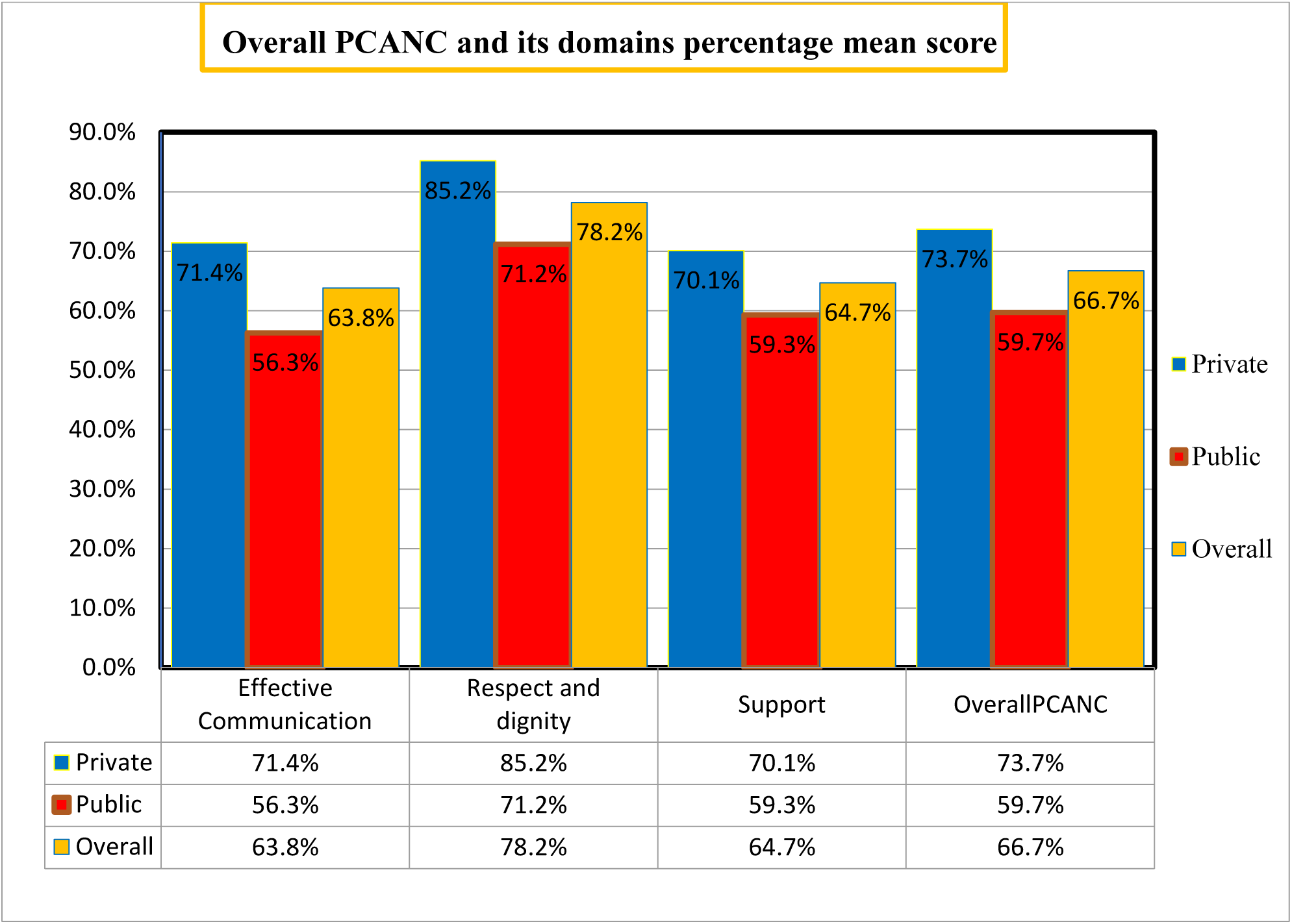
Percentage mean score of PCANC and its domains from the total expected score among pregnant women of Western Hararghe Zone, Oromia, Ethiopia, 2020.

