## Supplementary File 1 for "Levels of Person Centered Antenatal Care among Pregnant Women in Public and Private Health Facilities in Western Hararghe Zone, Ethiopia: An Institution Based Comparative Mixed Method Study Design"

**English Version Questionnaire**

| Respondents’ identification  001. Questionnaire Code______  002. Name of Health facility___________________ 003. Respondent’s address  004: Type of Health Facility  **Section 1: Socio-demographic characteristics of mother** | | |  |
| --- | --- | --- | --- |
| **S.N** | **Questions** | **Answers** | **Remark** |
| 101 | Mother’s age | __________yrs |  |
| 102 | What is your religion? | 1. Orthodox 2. Muslim 3. Protestant 4. Catholic 5. Other (specify)……… |  |
| 103 | Place of residence | 1. Urban 2. Rural |  |
| 104 | What is the highest grade you completed? | 1. Can’t read and write 2. Can read and write 3. Primary education (1-8) 4. Secondary education (9-12) 5. College/University |  |
| 105 | What is your Partner level of education? | 1. Can’t read write 2. Can read and write 3. Primary education (1-8) 4. Secondary education (9-12) 5. College/university |  |
| 106 | What is your present Occupation? | 1. House wife 2. Merchant 3. Government employee 4. Farmer 5. Daily work 6. Student 7. Other…….. |  |
| 107 | What is your present marital status? | 1. Single 2. Married 3. Divorced 4. Widowed |  |
| 108 | What is your ethnicity? | 1. Oromo 2. Amhara 3. Somale 4. Gurage 5. Others |  |
| 109 | What is your family monthly income? | __________birr |  |
| 110 | Self or household member work in health facility | 1. Yes 2. No |  |
| 111 | Have you experienced domestic violence? | 1. Yes 2. No |  |
| 112 | What is your participation in household decisions? | 1. Low participation 2. High participation |  |

**Section 2: level of Person Centered Antenatal Care**

**Effective communication Domain**

| 201 | During your time in the health, facility did the doctors, nurses, or other health-care providers introduce themselves to you when they first came to see you? | | 0. No, none of them 1. Yes, a few of them 2. Yes, most of them 3. Yes, all of them |
| --- | --- | --- | --- |
| 202 | Did the doctors, nurses, or other health-care providers call you by your name? | | 0. No, never 1. Yes, a few times 2. Yes, most of the time 3. Yes, all the time |
| 203 | Did the doctors, nurses, or other staff at the facility speak to you in a language you could understand? | | 0. No, never 1. Yes, a few times 2. Yes, most of the time 3. Yes, all the time |
| 204 | Were you told the results after you were weighed? | | 0. No, never 1. Yes, a few times 2. Yes, most of the time 3. Yes, all the time |
| 205 | Were you told the results after your blood pressure was taken? | | 0. No, never 1. Yes, a few times 2. Yes, most of the time 3. Yes, all the time |
| 206 | Were you told the results of the urine test? | | 0. No, never 1. Yes, a few times 2. Yes, most of the time 3. Yes, all the time |
| 207 | Were you told the results of the blood test? | | 0. No, never 1. Yes, a few times 2. Yes, most of the time 3. Yes, all the time |
| 208 | Were you told about the signs of pregnancy complications? | | 0. No 1. Yes |
| 209 | Were you told where to go if you had any complications? | | 0. No 1. Yes |
| 210 | Were you ever told what to expect in the course of your pregnancy and delivery | | 0. No 1. Yes |
| 211 | Were you talked about birth preparedness | | 0. No 1. Yes |
| 212 | Were you given advice on nutrition education | | 0. No 1. Yes |
| 213 | Were you given any information or counseled about breast feeding? | | 0. No 1. Yes |
| 214 | Did you feel you understood the purpose of any tests | | 0. No, never 1. Yes, a few times 2. Yes, most of the time 3. Yes, all the time |
| 215 | Understood the purpose of any medicines | | 0. No, never 1. Yes, a few times 2. Yes, most of the time 3. Yes, all the time |
| 2016 | Did you raise questions for health provider? | | 0. No, never 1. Yes, a few times 2. Yes, most of the time 3. Yes, all the time |
| 2017 | Did the doctors, nurses or other staff at the facility ask you if you had any questions | | 0. No, never 1. Yes, a few times 2. Yes, most of the time 3. Yes, all the time |
| 2018 | Did the doctors and nurses explain to you why they were doing examinations or procedures on you? | | 0. No, never 1. Yes, a few times 2. Yes, most of the time 3. Yes, all the time |
| 2019 | Did the doctors and nurses allow the position of your choice for your ANC examination? | | 0. No, never 1. Yes, a few times 2. Yes, most of the time 3. Yes, all the time |
|  | **Respect and dignity Domain** | | |
| 220 | | Did the doctors, nurses, or other staff at the facility treat you with respect? | 0. No, never 1. Yes, a few times 2. Yes, most of the time 3. Yes, all the time |
| 221 | | Did the doctors, nurses, and other staff at the facility treat you in a friendly manner? | 0. No, never 1. Yes, a few times 2. Yes, most of the time 3. Yes, all the time |
| 222 | | Did you feel the doctors, nurses, or other health-care providers shouted at you, scolded, insulted, threatened, or talked to you rudely? | 0. No, never 1. Yes, a few times 2. Yes, most of the time 3. Yes, all the time |
| 223 | | Did you feel like you were treated roughly like pushed, beaten, slapped, pinched, physically restrained, or gagged? | 0. No, never 1. Yes, a few times 2. Yes, most of the time 3. Yes, all the time |
| 224 | | Were you covered up with a cloth or blanket or screened with a curtain so that you did not feel exposed? | 0. No, never 1. Yes, a few times 2. Yes, most of the time 3. Yes, all the time |
| 225 | | Do you feel you could discuss your problems with health professionals without others not involved in your care | 0. No, never 1. Yes, a few times 2. Yes, most of the time 3. Yes, all the time |
|  | **Supportive care Domain** | |  |
| 226 | Did the doctors and nurses at the facility talk to you about how you were feeling? | | 0. No, never 1. Yes, a few times 2. Yes, most of the time 3. Yes, all the time |
| 227 | Did the doctors and nurses at the facility encourage male involvement? | | 0. No, never 1. Yes, a few times 2. Yes, most of the time 3. Yes, all the time |
| 228 | Did any staff at the facility ask you for kitu kidogo (unofficial cost)? | | 0. No, never 1. Yes, once 2. Yes, a few times 3. Yes, many time |
| 229 | Would you say you were treated differently because of any personal attribute | | 0. No, never 1. Yes, once 2. Yes, a few times 3. Yes, many time |
| 230 | Did you feel the health facility environment, including washrooms, latrine etc were clean? | | 0. No, never 1. Yes, a few times 2. Yes, most of the time 3. Yes, all the time |
| 231 | How did you feel about the amount of time you waited? What would you say? | | 0. Very short 1. Somewhat short 2. Somewhat long 3. Very long |
| 232 | Did you feel safe (calm and quiet) in the health facility? | | 0. No, never 1. Yes, a few times 2. Yes, most of the time 3. Yes, all the time |

**Qualitative part Tool**

**Observation checklist**

|  | **Interpersonal aspects of antenatal care observation check list** | Yes | No |
| --- | --- | --- | --- |
| 1. | Making women comfortable by providing seat and greeting |  |  |
| 2. | Interruption of women’s speech |  |  |
| 3. | Do health care workers treat clients respectfully |  |  |
| 4. | Does concerns of women asked about |  |  |
| 5. | Do health care workers explaining procedure to women |  |  |
| 6. | Do health care workers keep patient confidentiality or privacy |  |  |
| 7. | Do health care workers allocate sufficient time to each client |  |  |

**Thank You!!!**
